# Development and evaluation of the cooperative experiences measure

**DOI:** 10.64898/2026.05.21.26353820

**Authors:** Benjamin P. L. Meza, Ron D. Hays, Rebecca N. Dudovitz, Mitchell D. Wong

## Abstract

Given the growing crisis in youth mental health, there is a critical need to rebuild and sustain healthy social environments. Cooperative experiences (e.g., sports, clubs) may promote mental health but we lack rigorously tested measures to drive research and evaluation. This study sought to develop a measure of cooperative experiences and test associations with health.

We developed and revised a measure of cooperative experiences based on interdisciplinary literature and 20 cognitive interviews. We recruited youth aged 13-25 years (N = 262) through youth-serving organizations and snowball sampling to complete an investigator-administered (n = 50) or self-administered (n = 212) survey assessing cooperative experiences (48 items), mental and physical health, and demographics. We assessed item characteristics, dimensionality, reliability, and construct validity. Multivariable linear regressions were used to estimate the association between the total score and self-reported health.

Participants were 57% female, 69% Latino, 55% high school students, and 25% college students. The measure was reduced to 35 items (alpha = 0.90) with six subscales: sense of a unified group (7 items, alpha = 0.83), goal alignment (3 items, alpha = 0.80), inclusion and shared purpose (10 items, alpha = 0.88), social exclusion (2 items, alpha = 0.91), positive interdependence (7 items, alpha = 0.77), and negative interdependence (6 items, alpha = 0.87). A higher total score was associated with better self-reported mental health (beta = 0.25 standard deviation change in health score for each standard deviation change in cooperation scale, 95% CI [0.108, 0.394], p = 0.001) and self-reported general health (beta = 0.25, 95% CI [0.107, 0.395], p = 0.001).

The study provides preliminary support for the reliability and validity of a new measure of exposure to cooperative experiences among youth. The measure holds promise as a tool to examine the relationship between social environments and health outcomes in real-world settings.

## Introduction

Social environments for young people are changing in unprecedented ways as more of their peer interactions move online, and this shift is associated with an unprecedented global adolescent mental health crisis [1–3]. The shift has called into question our assumptions about what elements of social interactions are most important for adolescent development. In a hyper-connected world, superficial interactions (e.g., likes and subscription audiences) can displace face-to-face and more complex interactions with downstream negative effects on social skills (e.g., empathy, interpretation of facial/auditory cues) and prosocial bonding, further fueling disconnection and ill health [4–8]. We hypothesize that cooperative experiences may promote social and emotional skill building, positive peer connections, and mental health resilience.

Cooperation is defined as working interdependently with others toward shared goals and mutual benefit in a setting that creates a sense of inclusion [9,10]. There are three states of interdependence that may coexist to differing degrees within a single activity [11]. First, positive interdependence occurs when success can only be achieved as a group, not as individuals (e.g., a science project in which each student has a specific role that depends on other students completing their tasks, and the entire group is graded as a single unit). This aligns individual motivations and incentivizes mutually supportive peer interactions. Second, negative interdependence occurs when success is contingent on others’ failure (e.g., sprinting in track and field). Third, the absence of interdependence occurs when individuals work independently, and outcomes do not affect or depend on others (e.g., painting). Interdependence is not a continuous spectrum ranging from negative to positive. Rather an activity can have both negative and positive interdependence aspects. Negative interdependence can promote mastery, respect, and prestige, thereby enhancing social relationships in certain instances when social norms promote shared values (e.g., sportsmanship) over individual performance or when combined with positive interdependence (e.g., team-based competition) [12–15]. However, when negative interdependence is narrowly focused on individual success in a winner-takes-all social environment, individuals are incentivized to undermine social relationships to get ahead [13]. Although these states can coexist, highly cooperative experiences are expected to have a relative predominance of positive interdependence. In contrast, highly competitive experiences are expected to have a predominance of negative interdependence. Cooperative experiences are hypothesized to include additional elements such group identity, inclusive decision making, and belongingness to the group [17].

School-based interventions designed around cooperative elements have demonstrated salutary effects in multiple key domains, including academic achievement [18,19], social and emotional skills such as empathy and effective communication [20,21], peer social integration [21], and motivation due to a sense of belonging and shared purpose [20,22]. There is also promising evidence that these interventions may reduce internalizing (e.g., depressive symptoms) and externalizing (e.g., substance use) psychopathologies with possible mediation by social network mechanisms [23–25]. Exposure to cooperation is thought to improve health through at least two interconnected mechanisms: (1) building non-cognitive skills such as social awareness, empathy, and effective communication, and (2) promoting healthy social networks by increasing the formation of supportive relationships and increasing the influence of prosocial individuals in a community [26].

Despite growing evidence for specific use settings and classroom techniques, no measure of cooperation exists to study cooperation generally across contexts and to translate replicable principles beyond the classroom for the study of health and development in adolescents and young adults. The most developed research comes from the field of education, where several measures exist for assessing attitudes and acceptance of cooperative techniques in the classroom, as well as measures of fidelity that can be applied to online or offline exercises [27–32]. These measures are centrally focused on classroom environments and cooperation’s specific application to learning outcomes. In the business and management literature, two measures focus on entitativity (i.e., perception of being part of a unified group) or specific aspects of interdependence (e.g., task and reward interdependence), but they have not been extensively evaluated, are context-dependent, and are designed to examine interactions between coworkers [33,34]. One pre-post design study used a two-item measure of interdependence as a mediator to explain the beneficial changes seen after a particular sports event; however, very limited psychometric testing was performed [35]. Thus, we aimed to develop and test a measure of exposure to cooperative experiences that could be used to examine their role in life course health development and to evaluate cooperative interventions across a range of contexts. Uniquely, our measure was also designed with the expectation that different forms of interdependence are not mutually exclusive and thus may be measured simultaneously.

The current study is the first step in a multi-part, iterative process of scale development to create a measure of exposure to cooperative experiences and assess its psychometric properties. The goal is to develop a survey measure that can be used flexibly across a variety of contexts to assess the quality of cooperative experiences to which youth are exposed and to evaluate if such experiences are protective of health. The process is built upon an interdisciplinary literature, including pedagogy, psychology, sociology, and life-course health development research, to inform the creation of youth social environments and improve health and well-being.

## Materials and Methods

The measure development protocol consisted of four stages. In the first stage, we developed a draft measure based on the existing literature, comprising 66 items across 11 thematic domains. In the second phase, items were revised iteratively from cognitive interviews with 20 youth and research advisors from multiple disciplines. The resulting 54 items were administered by the principal investigator to 50 adolescents in the third stage. Finally, 48 items that were unchanged from the third stage were self-administered by 227 adolescents using an online platform (Research Electronic Data Capture, REDCap [36,37]).

### Stage 1

Creation of the initial inventory of items in Stage 1 took a deductive approach, retaining an inventory of items that was broad and inclusive of many domains that characterize cooperative activities, including: entitativity (qualities of being a group experience) [33], interdependence [11], attitudes and beliefs around cooperation [27,38], group membership [33], shared goals and purpose, adult and peer relationships, belongingness [39–41], internal motivations, intensity of exposure, and adult leadership. Item generation and review were performed in consultation with nine subject matter experts in developmental psychology, education, pediatrics, internal medicine, adolescent health, and measurement development.

Because evaluating all experiences of an individual was not feasible, the measure contains two parts: (1) an experience generator question that asks the respondent to nominate an impactful experience they have had (“Pick the one activity [in the last 12 months] you think has had the most impact on your life.”) and (2) questions about the characteristics of the nominated activity including: degree of impact, years of participation, entitativity, leadership, peer relationships, interdependence (overall, goals, rewards, tasks, resources, norms, decisions, identity), a sense of shared purpose and belonging felt as part of that activity. The measure was designed to capture the most impactful experience within a recall period as a sample of an individual’s overall exposure to cooperative experiences within that time frame. We used the recall period to calibrate the measure and better discriminate between youth with different frequencies of exposure to impactful experiences (i.e., a smaller time window decreased the pool of impactful experiences and thus permits better discrimination between youth with moderate to high frequency of exposure to impactful activities. We hypothesized that both the degree of experienced cooperation and frequency of exposure would drive the social network changes and engagement that influence health.

### Stage 2

Twenty adolescents and young adults aged 13-25 years participated in cognitive interviews to refine and vet the initial inventory of items based on face validity, age appropriateness, clarity, and understanding. Youth were eligible to participate if they were proficient in English and had not previously participated in the development of the measure. From 30 August 2023 to 25 October 2023, the study team recruited youth from public spaces and community events, and through 11 youth-serving community organizations, using a convenience sampling approach. We partnered with community organizations offering a broad range of youth programming, from highly cooperative to highly individualistic, including physical fitness activities, after-school academic support, health and wellness services, job training and placement, and social support services. Additional recruitment followed via snowball sampling, in which participants referred peers for eligibility screening and enrollment. Concurrently with youth cognitive interviews, the item inventory was reviewed by the subject matter experts.

Cognitive interviews informed revisions to the activity generator and individual items. In the original activities generator, participants nominated the most impactful experience in the last three years. Within this time interval, most youth could recall a long list of impactful experiences and ostensibly cooperative experiences were nearly universal. To improve our discrimination between youth with moderate to high frequency of impactful experiences and to reduce recall bias, we shortened the recall period to “in the last year.”

Cognitive interviews also informed the clarity and face validity of items and hypothesized domains. For example, the qualitative data suggested that some constructs, such as “attitudes and opinions of cooperation,” were related to cooperation but distinctly downstream and therefore were removed from the measure. Other constructs, such as “group size and composition,” were found to be highly variable and difficult to operationalize in a way that would be generalizable across activities and were more likely to modify the relationship between the measure and health indicators than to be components of the measure itself. Similarly, some of the original themes, such as “internal motivation,” were likely related to cooperation but were more appropriately classified as mediators and mechanistic constructs rather than elements of cooperative experiences themselves. Throughout Stage 2, new items were added, and old items were revised or eliminated, resulting in a net decrease from 66 to 54 items.

### Stage 3

In the third stage, recruitment of adolescents and young adults was expanded using the same recruitment strategy as in Stage 2. In this stage, the entire survey was administered to 50 participants. Recruitment occurred between 10 September 2024 and 20 December 2024. After completing the survey with their own responses, participants provided feedback on the survey as a whole and on specific items, focusing on clarity, acceptability, and administration feasibility. From these data, six items were eliminated because they either remained confusing to younger participants, were deemed overly complicated or burdensome, or provided little information, as all respondents answered nearly identically. Youth were also asked about their preferred survey administration mode, and the majority indicated that a self-administered online survey would be preferred. In Stage 3, no items were added or revised, so after vetting, the total number of items was 48.

### Stage 4

In the fourth stage, recruitment was again expanded by the same method, and 227 additional youth took the self-administered online survey version using REDCap [36,37]. Recruitment for stage 4 occurred from 17 July 2025 to 16 September 2025. Pooling the stage 3 and stage 4 survey responses yielded a sample of 277 adolescents and young adults. All but four items used Likert response options (*strongly agree, agree, not sure/uncertain, disagree, strongly disagree*). The remaining four items used a continuous slider scale from 0 to 10. Slider items were transformed to match the Likert scale (1 to 5) to avoid overweighting in the factor analysis and final measure scoring. Scale scores and the overall scale were scored as simple sums of item answer choices, with strongly agree, agree, not sure/uncertain, disagree, and strongly disagree receiving 2, 1, 0, -1, and -2 points, respectively. Slider items were scored in the same direction using a continuous score from 2 to -2. Two items (ex1 and ex2) were reverse coded for scoring (Table 2).

### Health measures

Health measures were collected as part of Stage 4 data collection. The primary health measure was self-rated general health as “How is your health in general? Would you say your health is…” *excellent, very good, good, fair, or poor* (scored 5 to 1 with higher values representing better health) [42,43]. Also measured was self-rated mental health, asked as “How would you rate your overall mental health?”) and reported using the same answer choices [44]; the Patient Health Questionnaire-2 (PHQ-2) [45], and the General Anxiety Disorder-2 score (GAD-2) [46]. The PHQ-2 included two questions, “Over the last 2 weeks, how often have you been bothered by the following problems?” (a) “Little interest or pleasure in doing things” and (b) “Feeling down, depressed or hopeless” each with four answer options: *not at all, several days, more than half the days, and nearly every day*. The GAD-2 was also asked in two questions, “Over the last two weeks, how often have you been bothered by the following problems?” (a) “Feeling nervous, anxious or on edge” and (b) “Not being able to stop or control worrying”, each with the same answer choices as the PHQ-2. PHQ-2 and GAD-2 were scored as a sum of the two questions (each scored from zero to 3, range of summed score 0-6).

### Covariates

Participants also reported their age, sex, racial/ethnic identity (multi-response item: American Indian or Alaska Native, Asian, Black or African American, Hispanic or Latino, Middle Eastern or North African, Native Hawaiian or Pacific Islander, White), student status (high school student, High school graduate/GED, college student (community college, university, college Graduate), and employment status (multi-response item: no job / not employed, employed part-time, employed full-time, self-employed). We did not ask respondents about gender-identity, sexuality, or identification with specific social groups.

### Analysis

All analyses were conducted in R using functions from the psych package for psychometric analyses [47,48]. Because most responses were collected via the self-administered survey, the study team first assessed the quality of the data and excluded survey responses that were 1) substantially incomplete (i.e., missing >50% of questions), 2) rushed (i.e., spending less than 2 seconds per question), or 3) found to have any non-sensical responses (e.g., non-sensical free text entry of the choice of activity, age of participation in activity older than current stated age). Because many items were specific to group activities, there was structured missingness when respondents nominated non-group activities. We analyzed item-level missingness, overall patterns of missingness, and missingness among respondents who nominated non-group activities. Next, we evaluated individual item characteristics, including summary statistics (means, standard deviations, distributions, skewness, kurtosis, inter-item correlations). Because types of activities and social roles often differ by sex/gender, item responses were stratified, and differences by sex were tested using Welch’s t-test to allow for unequal variances.

We conducted an exploratory factor analysis to determine the number of dimensions, the item set’s underlying structure, and whether items clustered into theoretically coherent domains across revisions. Exploratory factor analysis was performed using psych::fa() and minimal residual factoring (‘minres’) and oblique rotation (Promax) given that correlations between the factors were hypothesized. To account for structured missingness in the data, the study team used the full-information maximum-likelihood correlation matrix, calculated with psych::corFiml() under a missing-at-random (MAR) assumption, rather than the raw data. As in similar analyses [49], this approach permits the use of all available data despite structured item nonresponse, in this case, from the nomination of non-group activities. When missingness arises from logical skips, the missing values are expected to be non-informative (e.g., asking about competition between members of a group when the activity is not conducted in groups), making the missing-at-random assumption irrelevant to the analyses. Using this procedure requires no assumptions about data that are inconsistent with the observed data structure and enables analyses that leverage all available information. As the first step in a multi-step, iterative process, minimally sufficient model fit was defined as root mean square residual (RMSR) ≤ 0.08, Tucker-Lewis Index (TLI) > 0.8, root mean square error of approximation (RMSEA) ≤ 0.08, comparative fit index (CFI) ≥ 0.90, and standardized root mean square residual (SRMR) ≤ 0.08.

Identifying principal component eigenvalues greater than one, scree plot inspection, and parallel analyses were used to determine the number of dimensions. Items were iteratively removed based on multiple criteria including: low factor loadings (<0.4), cross-loading on multiple factors (loadings >0.3 with difference between factor loadings >0.2), low item-total correlations (< 0.3), redundancy (inter-item r >0.7), low communality (<0.2), high complexity (>2.5), and low corrected item-rest correlations (r drop < 0.3). Item skew was tolerated, as most respondents reported their most impactful experience in the last year as one of high cooperation, thereby skewing the underlying construct. After each iteration, an exploratory factor analysis was repeated on the reduced item set to confirm the factor structure’s stability. Internal consistency reliability was estimated with Cronbach’s alpha [50]. The final item set was chosen based upon theoretical interpretability; participant-reported face validity, clarity, and understanding; and statistical adequacy, while optimizing parsimony and recognizing the potential for future studies to perform further item revision.

To assess the relative contribution of total and subscales to the total score variance, we performed a variance decomposition. We then assessed for possible bifactor latent structure by comparing a single-factor confirmatory factor analysis (CFA) model to a bifactor CFA models, estimating using FIML maximum likelihood in the lavaan package. Model adequacy was summarized with the CFI, TLI, RMSEA, and SRMR. Finally, to quantify the relative contribution of the general factor and subscales, we computed the explained common variance (ECV) from standardized bifactor loadings.

We assessed construct validity by examining the statistical relationship between the measure of cooperative experiences and health measures. Health measures were collected in stage 4, so the sample size for these models was 212. First, bivariate models assessed associations between the total measure score and each individual measure dimension score for each health measure. Next models were adjusted for age, sex, Latino ethnicity, student status, and employment status as potential confounders of the relationship between cooperative experiences and health. Lastly, an interaction term between total score and sex was added to test modification by sex. Independent and dependent variables were standardized for interpretability across models. All statistical tests used a two-tailed alpha level of 0.05.

### Ethical considerations

All participants provided informed consent, and study procedures were approved by the institutional review board at the host institution (IRB-23-0917). In the first three stages, adult participants provided verbal consent. For minors, parental verbal permission was obtained along with verbal participant-informed assent. In Stage 4 (online survey), parental permission for minors was waived as approved by the ethics review, and written informed consent was obtained from all participants. Participants received monetary compensation for their participation.

## Results

### Sample characteristics

A total of 277 youth screened as eligible, consented, and participated in Stages 3 and 4. Sixteen surveys were excluded for being grossly incomplete, rushed, or having nonsensical responses as defined above. Demographic characteristics for the analytic set (N = 262) are shown in Table 1. Participant ages ranged from 13 to 25 years (median 17.0). Only four of the 262 nominated activities were mostly or entirely online.

**Table 1.** Participant characteristics (N = 262).

| **Table 1. Participant characteristics (N = 262)** |  |
| --- | --- |
| **Characteristic** | **n (%)** |
| Age, mean (SD) | 17.7 (2.56) |
| Female sex | 149 (56.9%) |
| Race/ethnicity |  |
| American Indian or Alaska Native | 0 (0%) |
| Asian | 35 (13.4%) |
| Black or African American | 15 (5.7%) |
| Hispanic or Latino | 166 (63.4%) |
| Middle Eastern or North African | 4 (1.5%) |
| Native Hawaiian or Pacific Islander | 0 (0%) |
| White, Non-Hispanic | 11 (4.2%) |
| More than one race/ethnicity | 22 (8.4%) |
| Prefer not to say / Missing | 9 (3.4%) |
| Student status |  |
| High school student | 144 (55.0%) |
| High school graduate/GED | 20 (7.6%) |
| College student | 65 (24.8%) |
| College graduate | 19 (7.3%) |
| Prefer not to say / Missing | 14 (5.4%) |
| Employment status |  |
| Not employed | 163 (62.2%) |
| Employed part-time | 64 (24.4%) |
| Employed full-time | 13 (5.0%) |
| Self-employed | 6 (2.3%) |
| Prefer not to say / Missing | 16 (6.1%) |
| Abbreviations: GED – General Education Development Degree.<br>Community college and university students are included in the<br>College student row. |  |

### Item characteristics and correlations

Table 2 reports descriptive statistics for all evaluated items and examines sex differences. Forty-seven items had ceiling effects *(strongly agree*) above 15%, and nine items had floor effects (*strongly disagree*) (S1 Fig). Many of the same items demonstrated high skew and kurtosis (Table 2). Only two items (ni3 and en3), both slider items, had significant differences by sex, although the absolute difference was less than 0.4. To evaluate redundancy among items, we calculated inter-item correlations. Figure 1 visualizes inter-item correlations with highly positive correlations in black and highly negative correlations in white (see S3 Table for complete correlation matrix). The item pairs with the largest correlation coefficients were ex1-ex2 (r = 0.83), in1-in3 (r = 0.77), ni4-ni5 (r = 0.73), and ni1-ni2 (r = 0.70).

**Table 2.** Item descriptive statistics (N = 262).

| Item | Question | Score |  | Distribution |  | Corr |
| --- | --- | --- | --- | --- | --- | --- |
|  |  | Mean | SD | Skew | Kurtosis | Item-rest |
| pi1 | My success in [activity] depended on how well my whole group did. | 2.40 | 1.27 | 0.55 | 2.06 | 0.37 |
| ot1 | Other participants needed help from me to do their part in [activity].<br>*Help could be things like support, information, special knowledge or skills. | 2.11 | 1.02 | 0.97 | 3.52 | 0.47 |
| pi2 | If important people saw the group doing well, they saw me doing well in [activity]. | 2.04 | 0.90 | 0.95 | 4.24 | 0.44 |
| pi3 | Each person's task mostly depended on others in [activity] doing their part. | 2.29 | 1.15 | 0.71 | 2.57 | 0.45 |
| ot2 | I needed the others in [activity] and they needed me to succeed. | 2.13 | 1.09 | 0.86 | 2.96 | 0.63 |
| pi4 | In the end, we were judged as a group/team instead of as individuals in [activity]. | 2.64 | 1.44 | 0.36 | 1.73 | 0.49 |
| pi5 | Almost everything I did in [activity] was to help my group reach its goal. | 1.86 | 0.90 | 0.94 | 3.17 | 0.44 |
| pi7 | I would give up what I want to help my group in [activity]. | 2.36 | 1.00 | 0.38 | 2.50 | 0.37 |
| ot3 | I needed help from other participants to do my part in [activity].<br>*Help could be things like support, information, special knowledge or skills. | 1.95 | 1.00 | 1.08 | 3.61 | 0.31 |
| en6 | Leaders in [activity] encouraged us to work together. | 1.67 | 0.87 | 1.65 | 6.15 | 0.52 |
| pi6 | It was important to me to respect decisions made by the group in [activity]. | 1.59 | 0.68 | 0.95 | 3.72 | 0.37 |
| ni1 | Leaders in [activity] encouraged us to compete against each other. | 3.26 | 1.39 | -0.21 | 1.69 | 0.36 |
| ni2 | People in the group/team competed against each other to see who was best in [activity]. | 3.13 | 1.44 | -0.12 | 1.62 | 0.34 |
| ni3 | How often did everyone compete against each other in [activity]? | 3.56 | 1.34 | -0.42 | 1.72 | 0.26 |
| ni4 | How often did you or your group compete against other groups in [activity]? | 3.31 | 1.51 | -0.25 | 1.52 | 0.45 |
| ni5 | A goal of [activity] was to win against other groups/teams of competitors. | 2.95 | 1.53 | -0.04 | 1.46 | 0.48 |
| ot4 | I needed to work on my own to do well in [activity]. | 2.11 | 1.12 | 0.73 | 2.47 | 0.11 |
| ot5 | How often did you do [activity] on your own WITHOUT working with others or competing with others? | 2.73 | 1.16 | 0.38 | 2.20 | -0.19 |
| ot6 | I felt happy to be part of [activity]. | 1.54 | 0.78 | 1.90 | 7.57 | 0.40 |
| in9 | I felt a sense of shared purpose in [activity]. | 1.44 | 0.68 | 1.83 | 7.28 | 0.35 |
| in6 | I felt safe in [activity]. | 1.59 | 0.77 | 1.53 | 5.64 | 0.41 |
| ot7 | I felt like I/we made progress toward my/our goal in [activity]. | 1.63 | 0.79 | 1.52 | 5.90 | 0.40 |
| in4 | I felt like an important member of [activity]. | 1.66 | 0.84 | 1.35 | 4.77 | 0.56 |
| in7 | I felt close to others in [activity]. | 1.71 | 0.88 | 1.33 | 4.38 | 0.57 |
| in10 | Being part of [activity] became a big part of what kind of people we were. | 1.99 | 0.97 | 1.08 | 4.02 | 0.38 |
| in8 | I was satisfied with how much I got to help make decisions in [activity]. | 1.93 | 0.91 | 0.89 | 3.41 | 0.43 |
| in1 | Everyone in [activity] treated each other fairly. | 1.81 | 0.96 | 1.16 | 3.66 | 0.36 |
| in3 | Everyone in [activity] made me feel wanted and accepted. | 1.76 | 0.90 | 1.18 | 3.95 | 0.45 |
|  |  | Mean | SD | Skew | Kurtosis | Item-rest |
| in2 | Others in [activity] listened to my opinions | 1.81 | 0.83 | 1.01 | 3.92 | 0.44 |
| ni6 | I felt like people were comparing me to others and judging me in [activity]. | 3.38 | 1.32 | -0.48 | 2.03 | 0.17 |
| ot8 | People in [activity] helped each other to achieve their group's goal. | 1.62 | 0.75 | 1.39 | 5.46 | 0.49 |
| in5 | I contributed to the choices made in [activity]. | 1.80 | 0.89 | 1.30 | 4.86 | 0.39 |
| en2 | In [activity], I felt like I was part of a team. | 1.75 | 1.03 | 1.48 | 4.43 | 0.58 |
| en1 | [activity] felt like a group activity to me. | 1.82 | 1.10 | 1.39 | 4.01 | 0.51 |
| en7 | If individual members disagreed with the group in [activity], they still followed the group's decision. | 2.31 | 0.96 | 0.56 | 2.72 | 0.29 |
| ot9 | If individual members disagreed with the group in [activity], they did what they wanted. | 3.46 | 1.17 | -0.45 | 2.19 | 0.01 |
| ga1 | My intentions were aligned with the overall group's intentions in [activity]. | 1.76 | 0.77 | 1.11 | 4.74 | 0.45 |
| ga3 | My personal goal worked well with the group's goal in [activity]. | 1.86 | 0.83 | 1.05 | 4.14 | 0.40 |
| ga2 | My personal values and the group/team values were mostly the same in [activity]. | 1.94 | 0.83 | 0.88 | 3.73 | 0.44 |
| ot10 | There was a common goal within my group/team in [activity], even if some of us may have started with different ideas. | 1.57 | 0.79 | 1.83 | 7.28 | 0.45 |
| ot11 | I was trying to accomplish the same thing as others in [activity]. | 1.82 | 0.92 | 1.24 | 4.32 | 0.19 |
| ot12 | If it was reasonable, I would sacrifice my time and energy to others in [activity]. | 2.10 | 0.92 | 0.87 | 3.73 | 0.40 |
| ot13 | In [activity], I knew the other people in my group. | 1.82 | 1.03 | 1.47 | 4.67 | 0.40 |
| en4 | There was a clear way to join a group/team. | 1.96 | 0.92 | 0.91 | 3.59 | 0.51 |
| en5 | There was a clear way to leave a group/team. | 2.38 | 1.13 | 0.70 | 2.85 | 0.36 |
| en3 | How often did everyone work together in [activity]? | 2.05 | 0.98 | 1.17 | 4.02 | 0.50 |
| ex1 | I felt isolated in [activity]. | 4.01 | 1.12 | -1.29 | 4.00 | -0.12 |
| ex2 | I felt left out in [activity]. | 4.07 | 1.02 | -1.27 | 4.29 | -0.10 |

**Fig 1.**
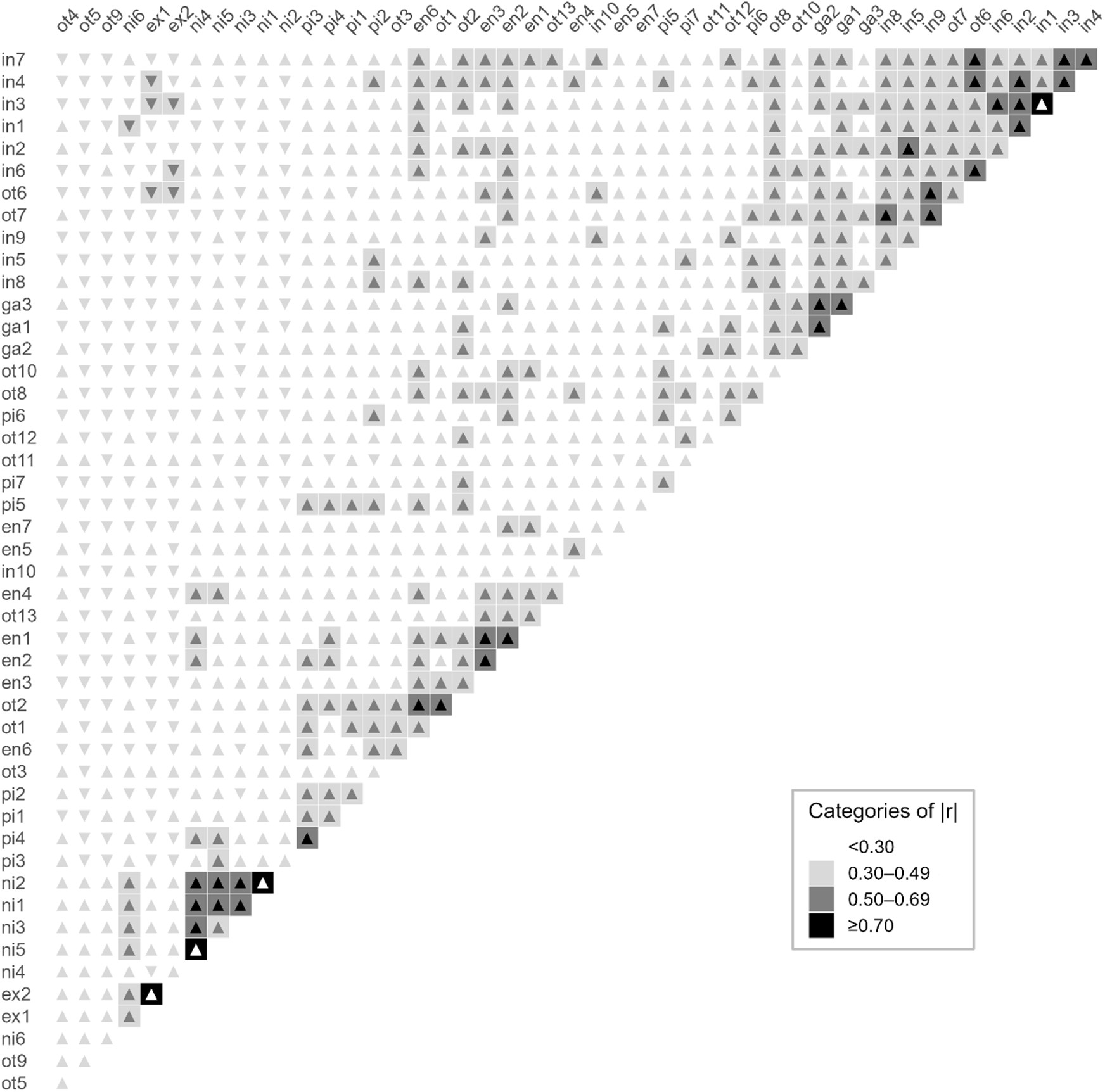
Inter-item correlation matrix heatmap. Each cell shows the Pearson correlation coefficient between two items. Fill color indicates the magnitude of the correlation’s absolute value (ranges from white <0.30 to black ≥ 0.70). Triangle orientation represents the correlation’s sign (▴ positive, ▾ negative). Items ordered to cluster higher item-item correlations. Numeric correlation matrix may be found in the supplement (S3 Table).

### Item-total correlations and internal consistency reliability

Table 2 also includes low item-rest correlations (<0.2) for ni1, ni2, ni3, ot4, ni6, and ot11. For the total score based on the original item set, Cronbach’s alpha was 0.92. In alpha-if-deleted analyses, no items were expected to improve internal consistency if removed.

### Construct Validity

Parallel analysis and the scree plot (S2 Fig) were consistent with an eight-factor structure. The exploratory factor analysis (Table 3) demonstrated minimally satisfactory model fit with RMSR = 0.04, TLI = 0.83, and RMSEA = 0.06. Rather than forming clearly bounded constructs, the item clusters showed overlapping thematic content, from which the following emerging factor themes were tentatively extrapolated: perception of shared benefits of positive interdependence and helping others in the group (F_i_1), negative interdependence (F_i_2), a sense of inclusion and shared purpose (F_i_3), perception of camaraderie and acceptance (F_i_4), self-definition of a group or team (F_i_4), shared goals and objectives (F_i_6), familiarity with others in the group (F_i_7), and social isolation (F_i_8). Five factors had inter-factor correlations >0.5 (Table 3). Nine items had low primary loadings (<0.4). The mean item complexity was 1.9. Six items had cross-loadings on more than one factor, and eighteen showed high complexity (> 2.0). In the initial factor analysis, three items demonstrated low communality (h^2^ <0.2; ot5, ot9, ot11).

**Table 3.** Standardized factor loadings, item communality, and item complexity of the initial item set.

| Item | F<sub>i1</sub> | F<sub>i2</sub> | F<sub>i3</sub> | F<sub>i4</sub> | F<sub>i5</sub> | F<sub>i6</sub> | F<sub>i7</sub> | F<sub>i8</sub> | $h^2$ | Complex |
| --- | --- | --- | --- | --- | --- | --- | --- | --- | --- | --- |
| pi1 | 0.87 |  | -0.34 |  |  |  |  |  | 0.61 | 1.38 |
| ot1 | 0.76 |  |  |  |  |  |  |  | 0.75 | 1.20 |
| pi2 | 0.70 |  |  |  |  |  |  |  | 0.56 | 1.16 |
| pi3 | 0.60 |  |  |  |  |  |  |  | 0.32 | 1.66 |
| ot2 | 0.56 |  |  |  |  |  |  |  | 0.60 | 1.60 |
| pi4 | 0.55 |  |  |  | 0.38 |  |  |  | 0.41 | 2.42 |
| pi5 | 0.51 |  |  |  |  |  |  |  | 0.53 | 1.44 |
| pi7 | 0.45 |  |  |  |  |  |  |  | 0.58 | 1.59 |
| ot3 | 0.42 |  |  |  |  |  |  |  | 0.66 | 2.22 |
| en6 | 0.33 |  |  |  |  |  |  |  | 0.67 | 2.99 |
| pi6 |  |  |  |  |  |  |  |  | 0.47 | 4.16 |
| ni1 |  | 0.81 |  |  |  |  |  |  | 0.63 | 1.05 |
| ni2 |  | 0.81 |  |  |  |  |  |  | 0.48 | 1.08 |
| ni3 |  | 0.81 |  |  |  |  |  |  | 0.55 | 1.07 |
| ni4 |  | 0.77 |  |  |  |  |  |  | 0.16 | 1.24 |
| ni5 |  | 0.71 |  |  |  |  |  |  | 0.46 | 1.42 |
| ot4 |  | 0.34 |  |  | -0.33 |  |  |  | 0.72 | 2.42 |
| ot5 |  |  |  |  |  |  |  |  | 0.41 | 4.50 |
| ot6 |  |  | 1.05† |  |  |  |  |  | 0.28 | 1.29 |
| in9 |  |  | 0.70 |  |  |  |  |  | 0.23 | 1.60 |
| in6 |  |  | 0.62 |  |  |  |  |  | 0.59 | 1.75 |
| ot7 |  |  | 0.51 |  |  | 0.36 |  |  | 0.21 | 2.10 |
| in4 |  |  | 0.50 |  |  |  |  |  | 0.53 | 1.68 |
| in7 |  |  | 0.43 |  |  |  | 0.30 |  | 0.45 | 2.38 |
| in10 |  |  | 0.31 |  |  |  | 0.30 |  | 0.58 | 2.21 |
| in8 |  |  |  |  |  |  |  |  | 0.42 | 4.04 |
| in1 |  |  |  | 0.94 |  |  |  |  | 0.49 | 1.08 |

| Item | F<sub>i1</sub> | F<sub>i2</sub> | F<sub>i3</sub> | F<sub>i4</sub> | F<sub>i5</sub> | F<sub>i6</sub> | F<sub>i7</sub> | F<sub>i8</sub> | h<sup>2</sup> | Complex |
| --- | --- | --- | --- | --- | --- | --- | --- | --- | --- | --- |
| in3 |  |  |  | 0.90 |  |  |  |  | 0.25 | 1.05 |
| in2 |  |  |  | 0.66 |  |  |  |  | 0.48 | 1.29 |
| ni6 |  | 0.37 |  | -0.40 |  |  |  |  | 0.24 | 3.11 |
| ot8 |  |  |  | 0.33 |  |  |  |  | 0.27 | 2.81 |
| in5 |  |  |  | 0.32 |  |  |  |  | 0.19 | 4.11 |
| en2 |  |  |  |  | 0.77 |  |  |  | 0.32 | 1.28 |
| en1 |  |  |  |  | 0.63 |  |  |  | 0.39 | 1.51 |
| en7 |  |  |  |  | 0.49 |  |  |  | 0.41 | 1.53 |
| ot9 |  |  |  |  | -0.31 |  |  |  | 0.56 | 2.99 |
| ga1 |  |  |  |  |  | 0.78 |  |  | 0.79 | 1.23 |
| ga3 |  |  |  |  |  | 0.73 |  |  | 0.59 | 1.45 |
| ga2 |  |  |  |  |  | 0.69 |  |  | 0.50 | 1.12 |
| ot10 |  |  |  |  |  | 0.41 |  |  | 0.74 | 2.22 |
| ot11 |  |  |  |  |  | 0.41 |  |  | 0.77 | 1.41 |
| ot12 |  |  |  |  |  |  |  |  | 0.59 | 2.31 |
| ot13 |  |  |  |  |  |  | 0.79 |  | 0.71 | 1.11 |
| en4 |  |  |  |  |  |  | 0.54 |  | 1.00 | 1.81 |
| en5 |  |  |  |  |  |  | 0.41 |  | 0.54 | 2.79 |
| en3 |  |  |  |  | 0.37 |  | 0.39 |  | 0.67 | 2.36 |
| ex1 |  |  |  |  |  |  |  | 1.02† | 0.65 | 1.09 |
| ex2 |  |  |  |  |  |  |  | 0.83 | 0.16 | 1.24 |
| Factor Correlations |  |  |  |  |  |  |  |  |  |  |
|  | F<sub>i1</sub> | F<sub>i2</sub> | F<sub>i3</sub> | F<sub>i4</sub> | F<sub>i5</sub> | F<sub>i6</sub> | F<sub>i7</sub> | F<sub>i8</sub> |  |  |
| F<sub>i1</sub> | 1.00 |  |  |  |  |  |  |  |  |  |
| F<sub>i2</sub> | 0.18 | 1.00 |  |  |  |  |  |  |  |  |
| F<sub>i3</sub> | 0.56 | -0.08 | 1.00 |  |  |  |  |  |  |  |
| F<sub>i4</sub> | 0.46 | -0.13 | 0.69 | 1.00 |  |  |  |  |  |  |
| F<sub>i5</sub> | 0.53 | 0.05 | 0.30 | 0.44 | 1.00 |  |  |  |  |  |
| F<sub>i6</sub> | 0.55 | 0.03 | 0.64 | 0.59 | 0.34 | 1.00 |  |  |  |  |
| F<sub>i7</sub> | 0.56 | 0.22 | 0.41 | 0.39 | 0.50 | 0.40 | 1.00 |  |  |  |
| F<sub>i8</sub> | 0.08 | 0.29 | -0.10 | -0.28 | -0.35 | -0.03 | -0.06 | 1.00 |  |  |
| Abbrev: h<sup>2</sup> = Communality; Complex = Complexity |  |  |  |  |  |  |  |  |  |  |
| † Values are pattern coefficients from maximum-likelihood exploratory factor analysis with promax rotation; pattern coefficients may exceed 1.00 (Heywood case) under oblique rotation and do not represent correlations. Items 47 and 48 were reverse coded for scoring. To preserve the interpretability of the factor's negative association, the inter-factor correlations shown correspond to the original coding. |  |  |  |  |  |  |  |  |  |  |

### Revised item set

The iterative process of measure refinement reduced the total number of items from 48 to 35 items. First, we dropped several items due to low Kaiser-Meyer-Olkin measures of sampling adequacy (< 0.6), including ot4, ot5, ot8, and ot9. Next, several items continued to demonstrate high cross loadings across two or more factors and so were iteratively dropped, including ot1, ot2, ot6, ot7, ot10, and ot13. With the number of cross-loaded items reduced, the resulting factor solution reduced from eight to six factors. Finally, items with weak factor loadings (i.e., ot3 and ot12) or low item-rest correlations and low communality (i.e., ot11) were dropped. Dropped items often had multiple criteria for which they were dropped. Several items were retained despite suboptimal statistical estimates because they represented important aspects of negative interdependence (i.e., ni1, ni2, ni3, ni6). This is hypothesized to be due to the complex coexistence of negative and positive interdependence in cooperative experiences, which are characterized by predominance of positive interdependence.

The final item set was consistent with a six-factor structure (Table 4) with minimally satisfactory model fit, RMSR = 0.04, TLI = 0.85, RMSEA = 0.062. Mean item complexity was 1.4 and no items cross-loaded. The six factors aligned with the following themes (S4 Table): (a) entitativity (i.e., perception of taking part in a unified group) and group norms (F_r_2, 7 items, alpha = 0.83); (b) goal alignment (F_r_6, 3 items, 0.80); (c) inclusion and shared purpose (F_r_1, 10 items, alpha = 0.88); (d) social exclusion (F_r_5, 2 items, alpha = 0.91); (e) positive interdependence (F_r_4, 7 items, alpha = 0.77); and (f) negative interdependence (F_r_3, 6 items, alpha = 0.87). Cronbach’s alpha of the total score for the whole item set was 0.90. Figure 2 provides a network visualization of the correlations between factors, demonstrating a tightly linked set of four factors: entitativity and group norms; goal alignment; inclusion and shared purpose; and positive interdependence. The total scale score was left-skewed with a median of 29.2 and a range from -21 to 67 (skew -1.2).

**Fig 2.**
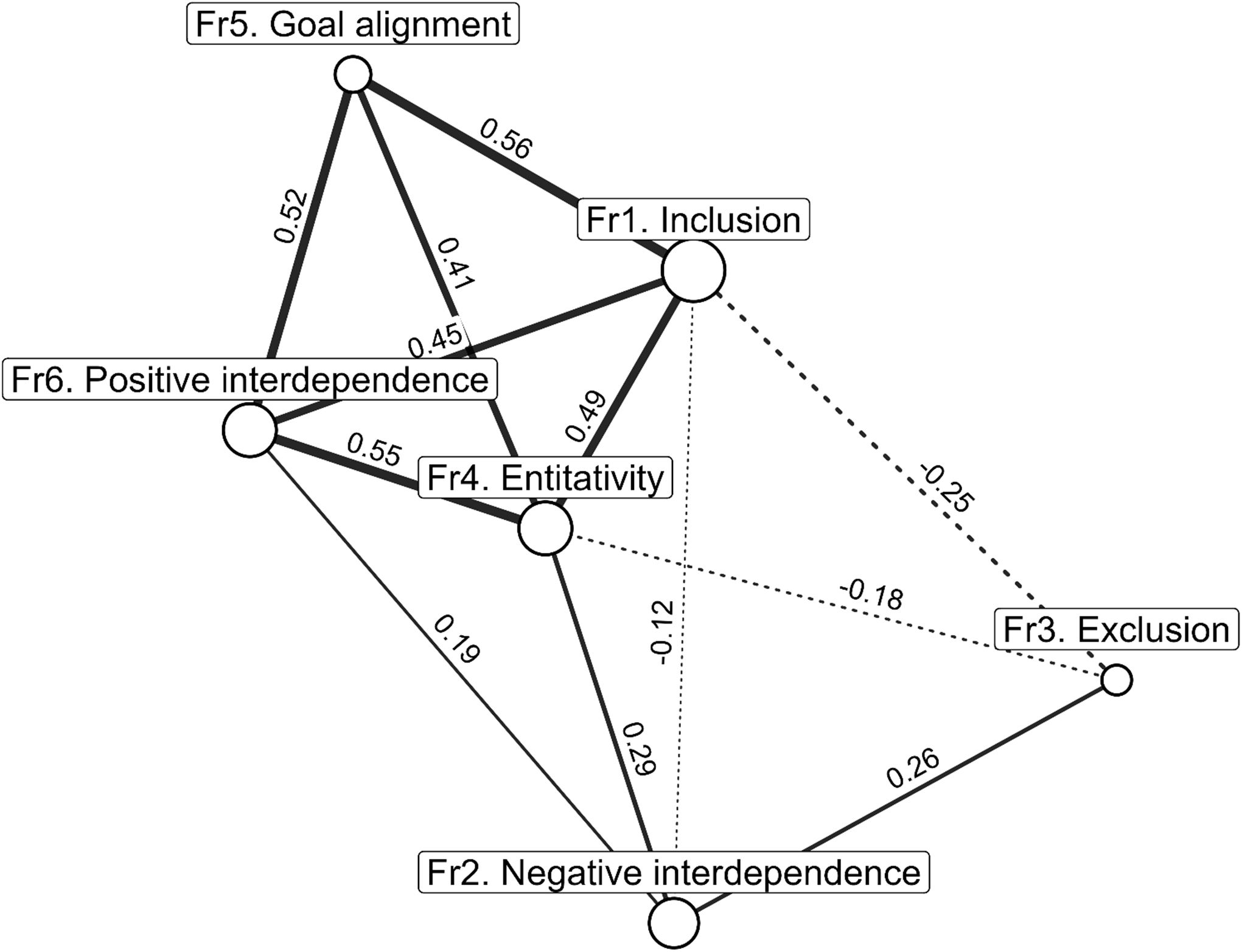
Network visualization of revised inter-factor correlations for the measure. Node size represents the number of items per subscale as listed in Table 4 (range 2-10). Lines represent the inter-factor correlation. Solid lines represent positive correlations while dotted lines represent negative correlations. Lines are labeled with the value of the inter-factor correlation. Line width is proportional to the absolute magnitude of the inter-factor correlation. Correlations < 0.10 are hidden for interpretability. Fr3 Exclusion items (ex1 and ex2) were reverse coded for scoring, however, to preserve the interpretability of the factor’s negative association, the inter-factor correlations shown correspond to the original coding. Layout based on Fruchterman-Reingold algorithm which pulls nodes with stronger inter-factor correlations closer together.

**Table 4.** Factor loadings, item communality, and item complexity of the revised item set. Subdomains: inclusion and shared purpose (F_r_1); entitativity (i.e., perception of taking part in a unified group and group norms (F_r_2); negative interdependence (F_r_3); positive interdependence (F_r_4); social exclusion (F_r_5); goal alignment (F_r_6).

| Item | F<sub>r</sub>1 | F<sub>r</sub>2 | F<sub>r</sub>3 | F<sub>r</sub>4 | F<sub>r</sub>5 | F<sub>r</sub>6 | h<sup>2</sup> | Complex |
| --- | --- | --- | --- | --- | --- | --- | --- | --- |
| in1 | 0.90 |  |  |  |  |  | 0.63 | 1.07 |
| in2 | 0.86 |  |  |  |  |  | 0.68 | 1.03 |
| in3 | 0.83 |  |  |  |  |  | 0.49 | 1.04 |
| in4 | 0.62 |  |  |  |  |  | 0.30 | 1.23 |
| in5 | 0.60 |  |  |  |  |  | 0.47 | 1.81 |
| in6 | 0.55 |  |  |  |  |  | 0.49 | 1.15 |
| in7 | 0.52 |  |  |  |  |  | 0.81 | 1.38 |

| Item | F<sub>r1</sub> | F<sub>r2</sub> | F<sub>r3</sub> | F<sub>r4</sub> | F<sub>r5</sub> | F<sub>r6</sub> | h<sup>2</sup> | Complex |
| --- | --- | --- | --- | --- | --- | --- | --- | --- |
| in8 | 0.42 |  |  |  |  |  | 0.67 | 2.06 |
| in9 | 0.42 |  |  |  |  |  | 0.43 | 1.52 |
| in10 | 0.34 |  |  |  |  |  | 0.64 | 1.63 |
| en1 |  | 0.82 |  |  |  |  | 0.31 | 1.29 |
| en2 |  | 0.78 |  |  |  |  | 0.73 | 1.03 |
| en3 |  | 0.68 |  |  |  |  | 0.40 | 1.06 |
| en4 |  | 0.67 |  |  |  |  | 0.22 | 1.17 |
| en5 |  | 0.48 |  |  |  |  | 0.48 | 2.15 |
| en6 |  | 0.47 |  |  |  |  | 0.50 | 1.68 |
| en7 |  | 0.38 |  |  |  |  | 0.56 | 1.72 |
| ni1 |  |  | 0.85 |  |  |  | 0.36 | 1.15 |
| ni2 |  |  | 0.85 |  |  |  | 0.49 | 1.07 |
| ni3 |  |  | 0.85 |  |  |  | 0.17 | 1.08 |
| ni4 |  |  | 0.75 |  |  |  | 0.23 | 1.23 |
| ni5 |  |  | 0.67 |  |  |  | 0.30 | 1.39 |
| ni6 |  |  | 0.40 |  |  |  | 0.41 | 2.96 |
| pi1 |  |  |  | 0.78 |  |  | 0.40 | 1.12 |
| pi2 |  |  |  | 0.69 |  |  | 0.47 | 1.43 |
| pi3 |  |  |  | 0.59 |  |  | 0.56 | 1.36 |
| pi4 |  |  |  | 0.59 |  |  | 0.37 | 1.67 |
| pi5 |  |  |  | 0.50 |  |  | 0.63 | 1.24 |
| pi6 |  |  |  | 0.35 |  |  | 0.67 | 2.14 |
| pi7 |  |  |  | 0.32 |  |  | 0.63 | 2.15 |
| ex1 |  |  |  |  | 0.93 |  | 0.78 | 1.02 |
| ex2 |  |  |  |  | 0.87 |  | 0.90 | 1.06 |
| ga1 |  |  |  |  |  | 0.91 | 0.50 | 1.11 |
| ga2 |  |  |  |  |  | 0.64 | 0.66 | 1.27 |
| ga3 |  |  |  |  |  | 0.64 | 0.64 | 1.17 |
| **Factor Correlations** |  |  |  |  |  |  |  |  |
|  | F<sub>r1</sub> | F<sub>r2</sub> | F<sub>r3</sub> | F<sub>r4</sub> | F<sub>r5</sub> | F<sub>r6</sub> |  |  |
| F<sub>r1</sub> | 1.00 |  |  |  |  |  |  |  |
| F<sub>r2</sub> | 0.53 | 1.00 |  |  |  |  |  |  |
| F<sub>r3</sub> | -0.13 | 0.31 | 1.00 |  |  |  |  |  |
| F<sub>r4</sub> | 0.50 | 0.64 | 0.22 | 1.00 |  |  |  |  |
| F<sub>r5</sub> | -0.28 | -0.19 | 0.27 | 0.00 | 1.00 |  |  |  |
| F<sub>r6</sub> | 0.62 | 0.48 | -0.03 | 0.57 | -0.06 | 1.00 |  |  |

Abbrev: h<sup>2</sup> = Communality; Complex = Complexity

Variance decomposition of the observed total score demonstrated that four factors accounted for a large proportion of the total variance: inclusion and shared purpose (0.25), entitativity (0.23), positive interdependence (0.22), negative interdependence (0.18), goal alignment (0.08), and exclusion (0.04). In assessing the relative contributions of a general factor and subscales, we found the single-factor CFA to have poor fit (CFI = 0.43, TLI = 0.40, RMSEA = 0.12, SRMR = 0.14). A bifactor CFA model fit the data much better (CFI = 0.86, TLI = 0.84, RMSEA = 0.06, SRMR = 0.09). The general factor accounted for a substantial proportion of the latent factor common variance (ECV = 0.42), indicating that the total score provided meaningful information to measure interpretation while substantial variation remained specific to the subscales. Taken together, these results supported the use of a total score alongside domain-specific factors.

### Self-Rated Health

Average self-reported mental health was 3.45 (SD 1.15), general health was 4.22 (SD 0.73), total PHQ-2 was 1.06 (SD 1.41), and total GAD-2 was 1.38 (SD 1.62). In multivariable models with standardized independent and dependent variables (Fig 3), self-reported mental health was positively associated with the total score (β = 0.25, 95% CI [0.11, 0.39], p < 0.001), Inclusion (β = 0.25, 95% CI [0.11, 0.38], p < 0.001), positive interdependence (β = 0.15, 95% CI [0.01, 0.30], p = 0.040), and negative interdependence (β = 0.16, 95% CI [0.01, 0.30], p = 0.034). Associations with entitativity (β = 0.12, 95% CI [–0.02, 0.27], p = 0.093), goal alignment (β = 0.13, 95% CI [–0.01, 0.28], p = 0.078), and social exclusion (β = 0.04, 95% CI [–0.10, 0.18], p = 0.581) were not statistically significant.

**Fig 3.**
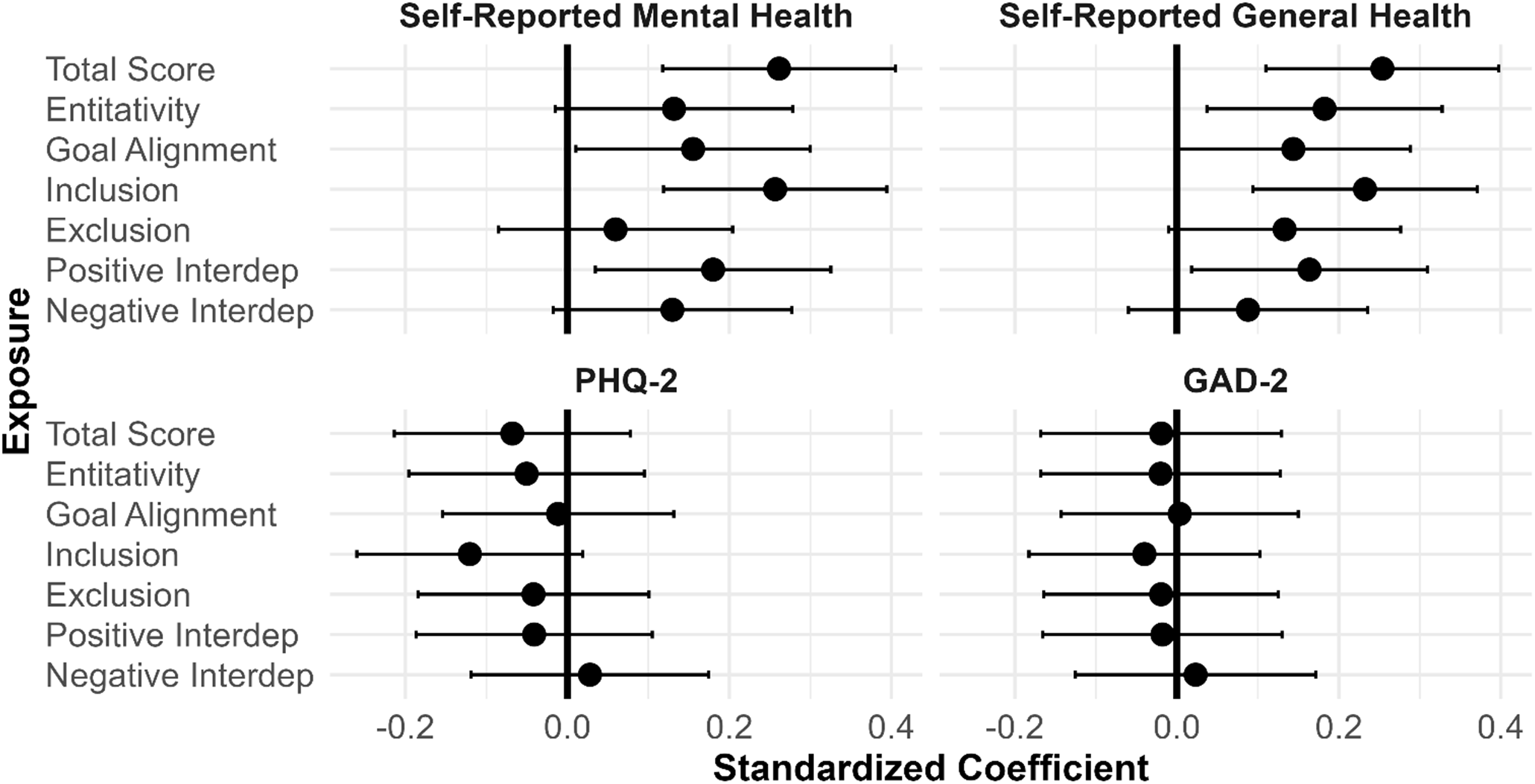
Associations between the cooperative experiences score (total score and individual dimensions) and health indicators. Each graph represents a separately modeled health indicator (self-reported mental health, self-reported health, PHQ-2, GAD-2). Measure scores (total; entitativity; goal alignment; inclusion and shared purpose; social exclusion; positive interdependence; negative interdependence subscales) are listed on the vertical axis. Circles represent standardized beta coefficients for each model (standard deviation change for each standard deviation change in independent variable) and horizontal brackets represent 95% confidence intervals around the estimate. Models were adjusted for age, sex, Latino ethnicity, student status, and employment status.

Self-reported general health was also positively associated with the total score (β = 0.25, 95% CI [0.11, 0.40], p < 0.001), inclusion (β = 0.23, 95% CI [0.09, 0.37], p = 0.001), and entitativity (β = 0.18, 95% CI [0.03, 0.32], p = 0.016). Positive interdependence (β = 0.15, 95% CI [0.00, 0.29], p = 0.051), goal alignment (β = 0.13, 95% CI [–0.01, 0.28], p = 0.077), negative interdependence (β = 0.11, p = 0.151), and social exclusion (β = 0.12, p = 0.094) were not significantly associated with general health.

PHQ-2 and GAD-2 total scores were not significantly associated with the total score or any subscale before accounting for modification by sex (all p > 0.05).

In interaction models, sex did not appear to moderate the association between cooperative experiences and self-rated health or self-rated mental health. In contrast, the interaction terms for cooperative experiences among females were significant for both depressive symptoms and anxiety symptoms (PHQ-2: β = -0.32, 95% CI [-0.60, -0.04], p = 0.025; GAD-2: β = -0.32, 95% CI [-0.60, -0.04], p = 0.026). Although depressive symptoms (PHQ-2) and anxiety symptoms (GAD-2) were not significantly associated with the exposure for males (PHQ-2: β = 0.13, 95% CI [-0.09, 0.35], p = 0.252; GAD-2: β = 0.18, 95% CI [-0.04, 0.40], p = 0.116), the linear combination for females was negative for both outcomes (PHQ-2: β = -0.19; GAD-2: β = -0.14), suggesting that greater exposure may be protective for depressive and anxiety symptoms among females. The main term for sex was not significant for any outcome (all p > 0.34). When subscales were modeled separately, sex appeared to moderate the association between three of the subscale scores and health outcomes: entitativity and group norms; inclusion and shared purpose; and social exclusion (S5 Fig).

## Discussion

This study presents initial development and evaluation of a measure of cooperative experiences. A literature review, cognitive interviews, and consultation with experts in the field provided initial face validity and improved clarity, understanding, and comprehensiveness. Piloting the inventory of items with youth aged 13 to 25 years yielded evidence of reliability, construct validity, and hypothesized associations with self-reported mental and general health as well as sex-specific associations with PHQ-2 and GAD-2. The factor analysis identified six subscales: entitativity and group norms; goal alignment; inclusion, and shared purpose; social exclusion; positive interdependence; and negative interdependence. These findings lay the groundwork for future research to (1) further evaluate the refined measure and (2) apply the tool to address important health questions about how social spaces, from school environments to extra-curricular activities and online platforms, may be modified to be more cooperative and thus, perhaps, promote better mental and physical health.

Iterative revisions produced a revised 35-item measure characterized by six subscales and acceptable internal reliability and model fit. Parallel analysis, scree plot, and exploratory factor analysis of the original 48-item set suggested an 8-factor model but also revealed redundancy, multiple items cross-loading or weakly loading, and items with low correlations with the construct. The revised 35-item measure demonstrated greater face validity and clearer factor structure, while improving the interpretability of the total scale and its subscales. A qualitative network analysis of the inter-factor correlations suggests that four subscales cluster closely (entitativity and group norms; goal alignment; inclusion and shared purpose; and positive interdependence) while two factors remain more peripheral (negative interdependence and social exclusion). Entitativity and group norms serve as a bridge: inclusion is negatively correlated with exclusion, and positive interdependence is positively correlated with negative interdependence. Interestingly, the data also showed that when asked to nominate impactful experiences in their recent past, adolescents tended to nominate activities higher in cooperation (i.e., the distribution was left-skewed) and rarely an entirely online activity. This finding supports the importance of social engagement in adolescence and, more specifically, that adolescents themselves find in-person cooperative activities valuable.

Teamwork is a latent construct closely related to cooperation that has become increasingly salient in biomedical literature. While there is no single consensus measure of teamwork, the field derives from an effort to better understand what defines a “team” and most measures focus on quality of communication (e.g., clear leadership, shared mental models) and synchronization of task completion (e.g., adaptability, effective coordination) [51,52]. While we have defined and sought to validate exposure to cooperative experiences as a social process with important implications for youth health development, teamwork is most often evaluated in relationship to productivity, particularly in the workplace (e.g., quality of product, cost savings) [51,52]. Of the much more limited number of teamwork measures in adolescent populations the Teamwork Scale for Youth (TSY) examines attitudes and perceptions of teamwork (e.g., “I think teamwork is important”) and is associated with improved social competencies (e.g., ability to make friends), but we were unable to identify any such measures that examined health outcomes [53]. Although we have focused less on communication and task completion, the findings of this study suggest conceptual alignment with measures of teamwork that include structural (e.g., positive interdependence) as well as relational (e.g., psychological safety) dimensions [52]. Whether high-quality teamwork may enhance the potential health benefits of cooperative experiences remains to be tested. Future analyses may formally test the convergent validity of this measure of cooperative experiences with comparable measures of teamwork.

Among the findings, the differential associations among female adolescents have important implications. Adolescent girls show a more direct reliance on supportive peer relationships for self-esteem and mental health, supporting the hypothesis that cooperative experiences may have specific benefits for girls [54–56]. Because social connections differently support mental health resilience for adolescent girls compared to boys, we described differences in item responses by sex and examined modification by sex in our health indicator models. Interestingly, we found only a few meaningful differences in item responses by sex. A more in-depth analysis may examine types of activities nominated by sex/gender and whether certain highly cooperative activities are more commonly nominated by certain groups. We found no modification by sex of the relationship between cooperative experiences and self-reported mental or general health. Before accounting for modification by sex, we found no association between exposure to cooperative experiences and PHQ-2 or GAD-2. However, interaction models showed significant modification by sex and demonstrated a protective relationship for depression and anxiety among female adolescents that males did not share. Modification appeared most pronounced for three subscales (entitativity and group norms; inclusion and shared purpose; and social exclusion). The current mental health crisis has disproportionately impacted adolescent girls [1], making the potential benefits of cooperative experiences even more important to investigate further.

These analyses inform continued revision of the measure in an iterative fashion. Most items showed ceiling effects or skew, limiting discrimination at the upper end of the cooperation scale. Future iterations will adjust answer choices to better accommodate the upper end of the response range (e.g., always, often, sometimes, rarely, never) and rephrase items to increase their difficulty (e.g., “I *always* felt like I was part of a team”). Improvements may also result from reducing the recall period, as was done following cognitive interviews. A shorter recall period (e.g., most impactful activity in the last month vs. the last 12 months) may help optimize sampling of impactful experiences to better differentiate youth with more frequent exposure from those with less frequent exposure. Some of the negative interdependence items (ni1, ni2, ni3 and ni6) demonstrated either mediocre psychometric properties or high item-item correlations in their current form. In lieu of discarding these questions, future modifications can attempt to capture more nuance. For example, some of the mediocre psychometric properties may be due in part to situations in which positive and negative interdependence occur at different levels of the activity or may even be in conflict (e.g., an employee may be incentivized to work well with colleagues to complete a task but derive personal benefit in the form of promotion if they can outperform peers). Using more precise language that allows the two forms of interdependence to coexist may improve clarity for the respondent and stronger psychometric properties for the items. For example, instead of asking “How often did people try to win or do better than each other in [activity]?” the question could be prefaced with “When you weren’t working together, how often did you try to win or do better than each other in [activity]?” In this study, we retain these items despite possible negative effects on parsimony and the TLI. Yet future studies with new data may find that further item reduction is possible without compromising construct validity.

The study’s findings should be interpreted considering the following strengths and limitations. This study benefited from a community-based sample of individuals with diverse racial/ethnic, educational, and employment backgrounds. Next, the four-stage design provided multiple opportunities to incorporate both qualitative and quantitative data into the measure development process, including feasibility and acceptability testing in investigator-administered and self-administered online formats and across a broad age range. The primary limitations of the study include the cross-sectional design, which does not permit causal or temporal analysis, and the small and demographically skewed study sample, which limits generalizability. There is some evidence that cooperative experiences may have differential effects or different mechanisms of effect in more collective cultures [57]. Future studies should use longitudinal data collection and temporal analysis, along with measures of cultural values, to better assess the performance of the measure over time and its relationship to health outcomes. Respondents were only asked about their sex but cooperation is fundamentally a social process so future studies would benefit from examining gender and specific social groups. Both independent and dependent variables were self-reports, which is optimal for some measures such as mental health, but observed measures of cooperation from teachers or coaches and objective measures of health would improve internal validity and the hypothesized connection to health. Lastly, this study cannot account for potential selection bias into cooperative experiences by individual personality or contextual factors that may also impact health. To our knowledge, no research to date has examined the predictors of engagement in cooperative activities, an important next step for developing a causal model and interventions that may be derived from this line of investigation.

## Conclusions

At a historical moment when more of our social interactions are through online platforms than ever before and youth are feeling both disconnected and mentally distressed, cooperative experiences are a promising protective factor for further exploration. This study reports the first psychometric analysis of a 35-item measure of cooperative experiences in this multi-part, iterative process of measure development. The analysis identified six subdomains supported by construct validity, reliability, and cross-sectional associations with self-reported health indicators. The study findings and the development of the measure are a promising first step toward connecting two disparate areas of research to improve health. Educators have amassed substantial empirical knowledge of innovative methods for fostering cooperation in the classroom [58], and health researchers have identified that this new generation of hyper-online youth reports historic levels of social isolation and mental distress [1]. Evidence from the classroom also suggests that when youth are exposed to cooperative activities, they develop favorable attitudes about cooperation and may be more likely to engage further, setting up a virtuous feedback loop [31]. Future studies may incorporate temporal analyses, test more parsimonious item sets, model relationships between subscales and potential mechanistic mediators (e.g., social-emotional skills, social network connectivity), and study the validity of these measures in larger, more diverse samples (e.g., greater cultural diversity). The measure under development has the potential to enable both the epidemiological study of cooperation as a protective factor for health and may be applied to evaluate interventions to build or adapt existing social spaces, both online and off. Lastly, technology-enabled measures of social interactions are rapidly growing across academia and commercial online platforms. A deeper understanding of the health impacts of social organizational structures – such as cooperation – across the life course could enable more accurate evaluation and prediction of how design choices by these actors could influence future health outcomes.

## Data Availability

Data cannot be shared publicly because they were collected from minors. Data are available from the UCLA Dataverse to qualified researchers who meet the criteria for accessing confidential data.

## Acknowledgments

None.

## Supporting information

**S1 Fig.**
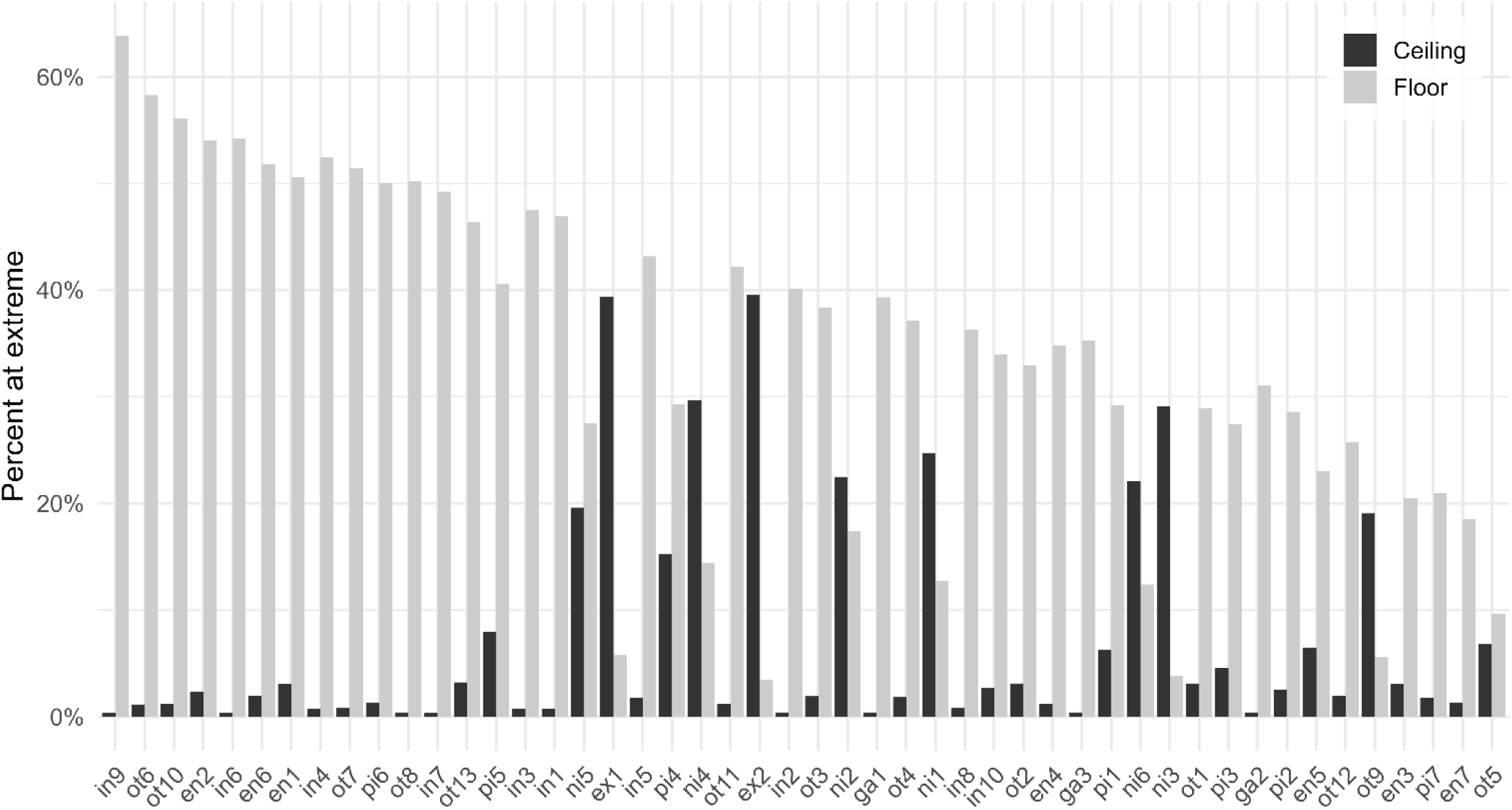
Floor and ceiling effects by item. The percent of responses (vertical axis) at the extreme of the response range for each item (horizontal axis). Grey indicates the percentage of responses at the ceiling and black indicates the percentage of responses at the floor of the response range.

**S2 Fig.**
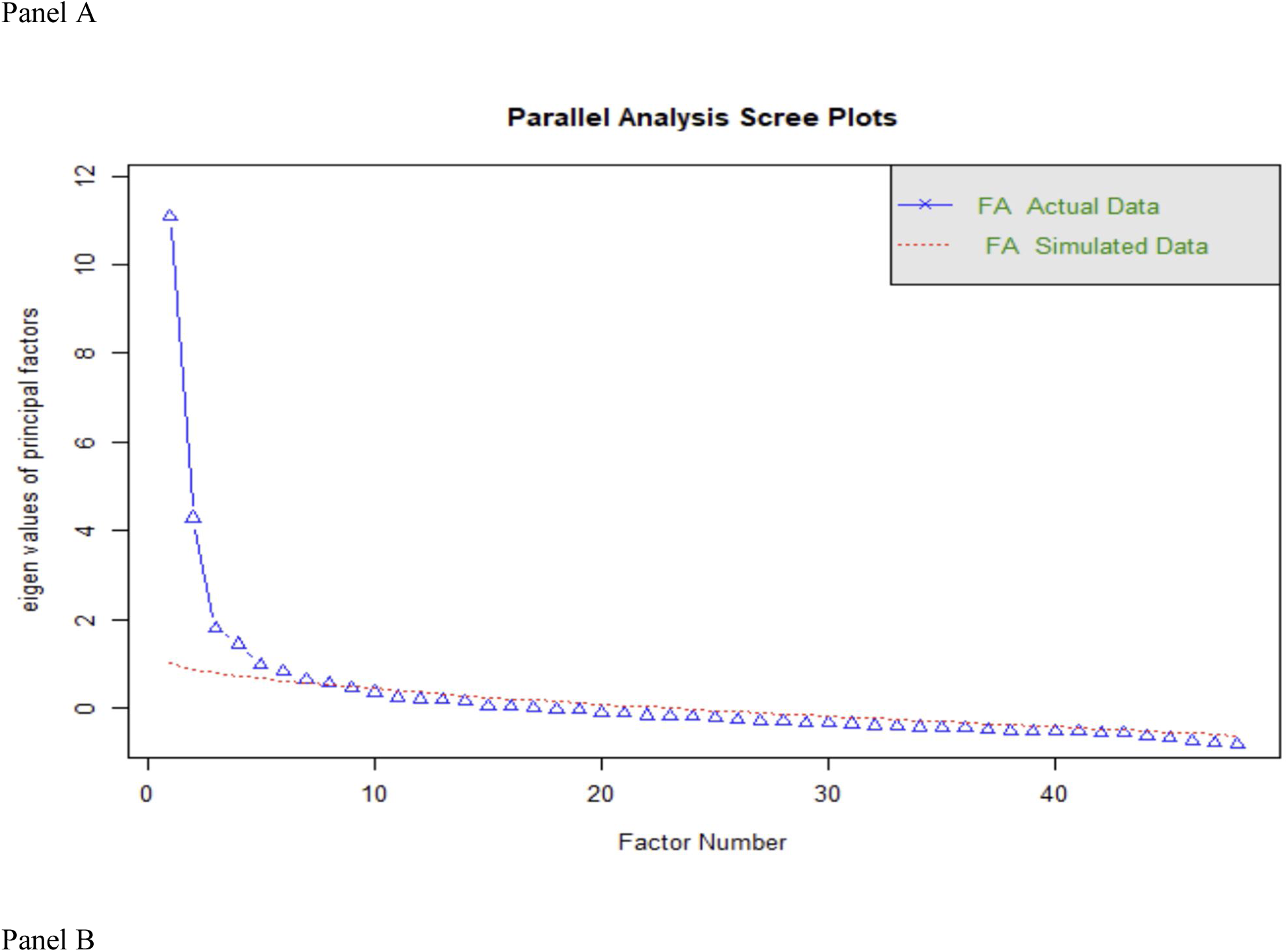
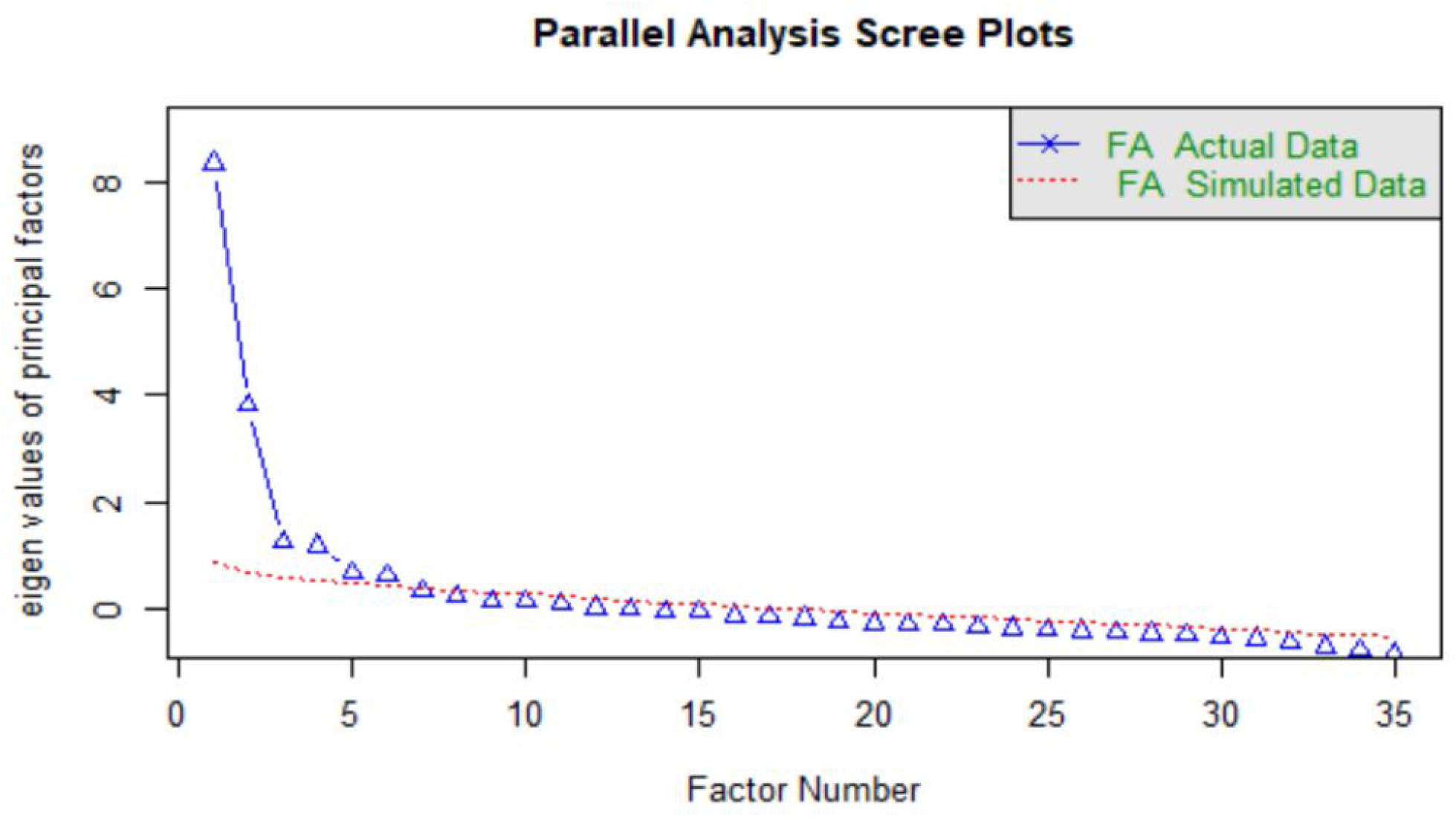
Scree plots. Scree plots for the original item set (Panel A) and the revised item set (Panel B).

**S3 Table.**
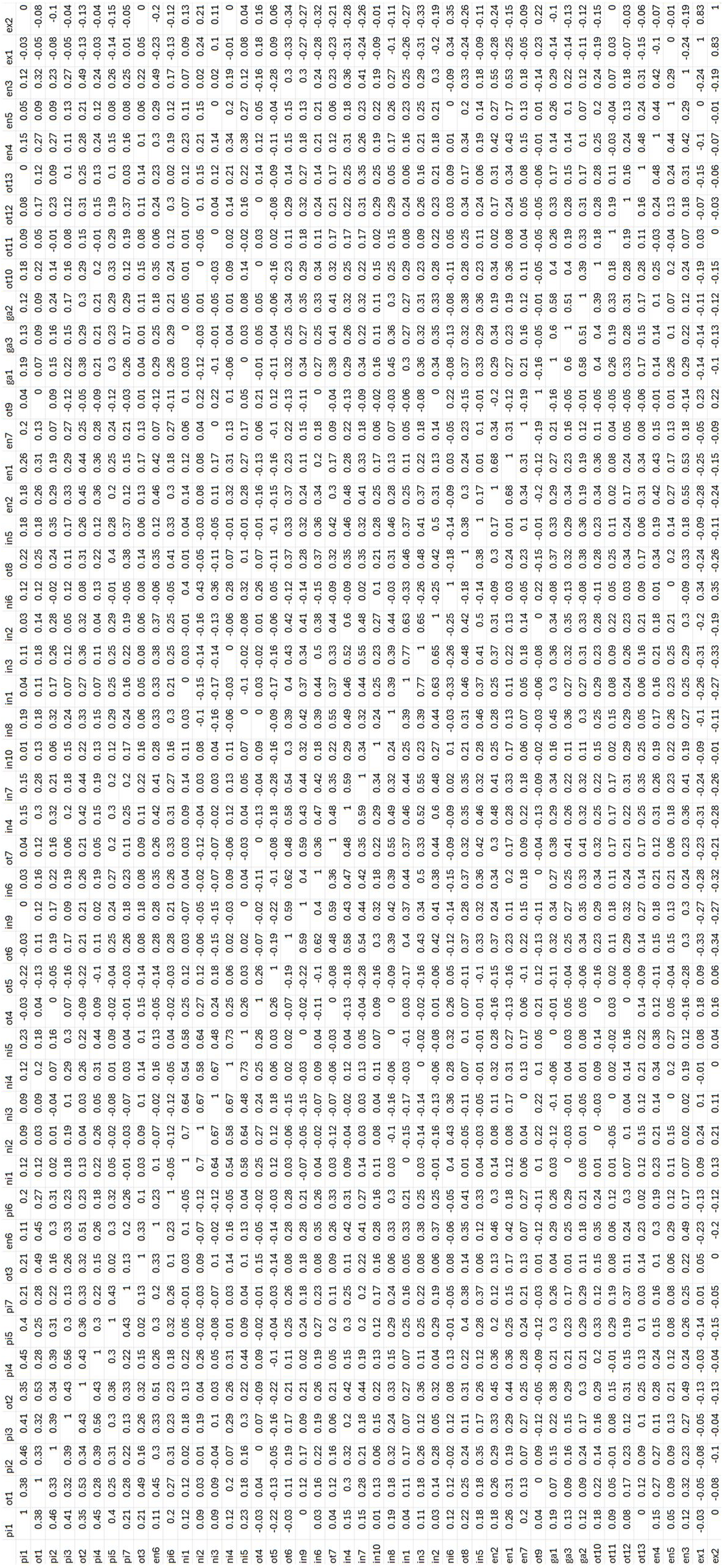
Item-item correlations.

**S4 Table.**
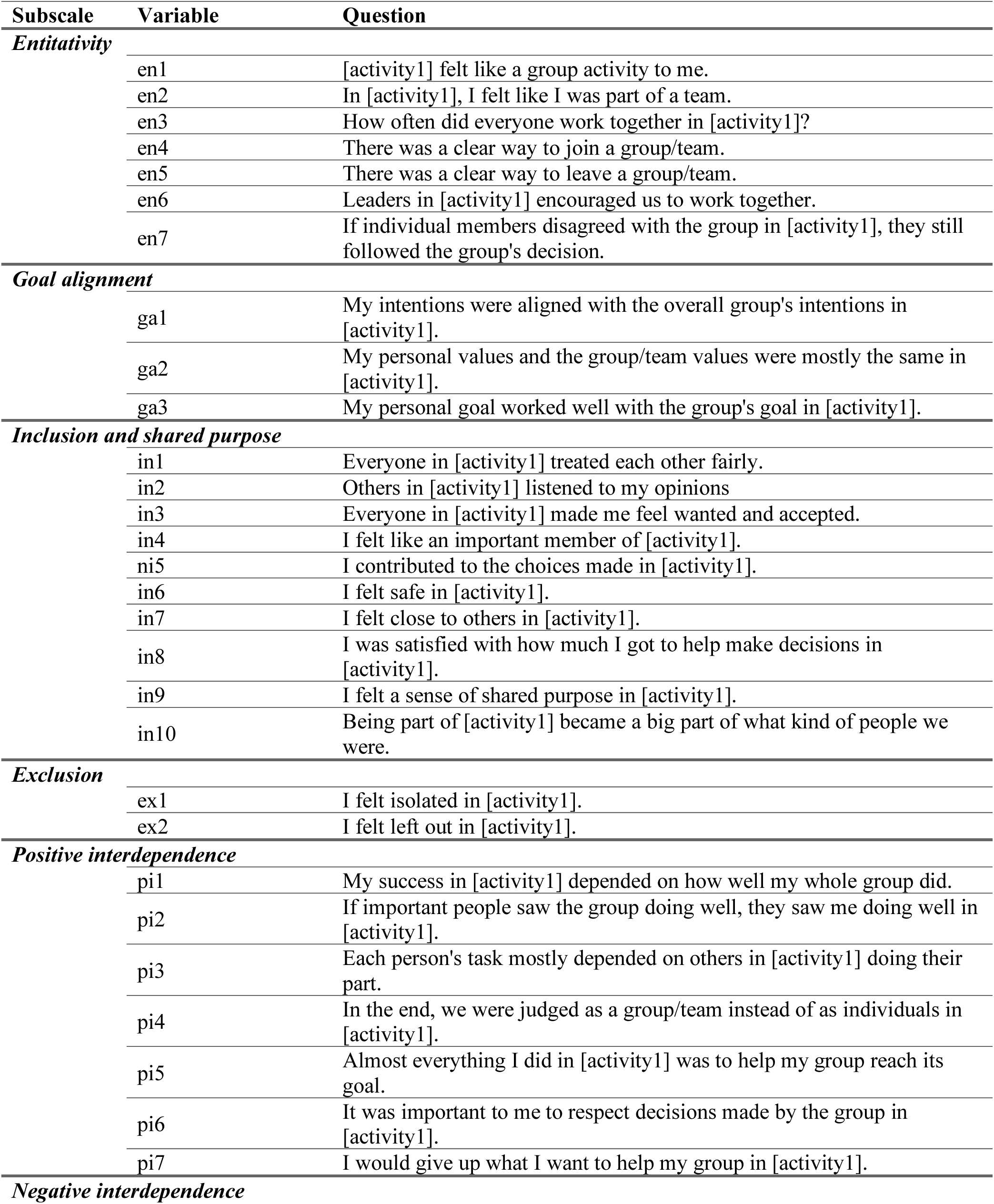
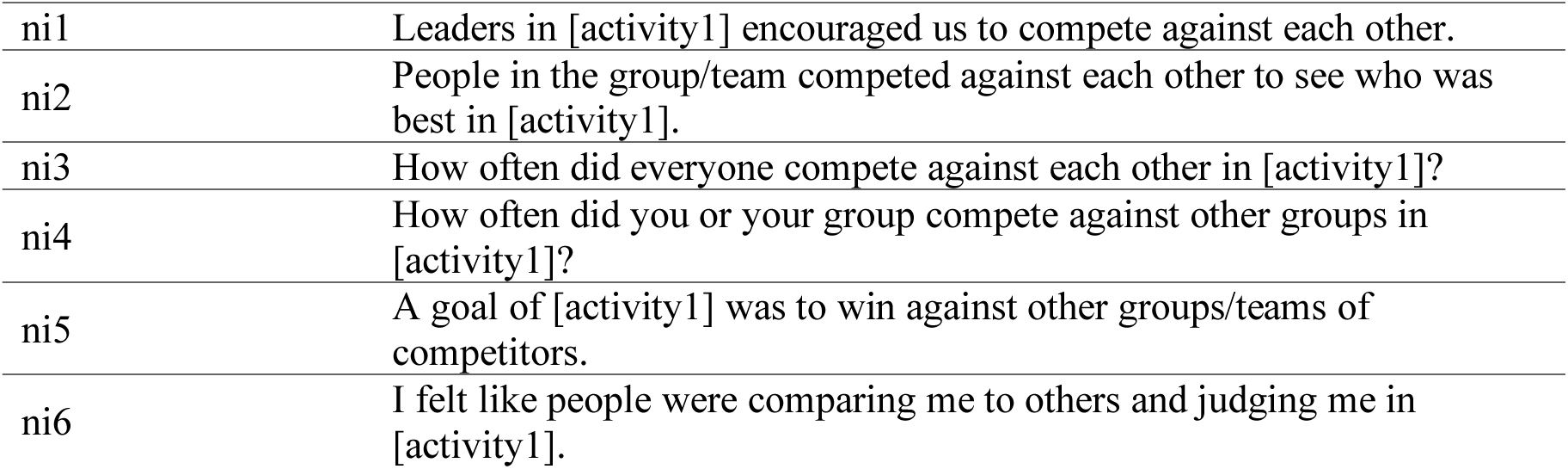
List of items organized by subscale in the revised measure.

| Subscale | Variable | Question |
| --- | --- | --- |
| ***Entitativity*** |  |  |
|  | en1 | [activity1] felt like a group activity to me. |
|  | en2 | In [activity1], I felt like I was part of a team. |
|  | en3 | How often did everyone work together in [activity1]? |
|  | en4 | There was a clear way to join a group/team. |
|  | en5 | There was a clear way to leave a group/team. |
|  | en6 | Leaders in [activity1] encouraged us to work together. |
|  | en7 | If individual members disagreed with the group in [activity1], they still followed the group's decision. |
| ***Goal alignment*** |  |  |
|  | ga1 | My intentions were aligned with the overall group's intentions in [activity1]. |
|  | ga2 | My personal values and the group/team values were mostly the same in [activity1]. |
|  | ga3 | My personal goal worked well with the group's goal in [activity1]. |
| ***Inclusion and shared purpose*** |  |  |
|  | in1 | Everyone in [activity1] treated each other fairly. |
|  | in2 | Others in [activity1] listened to my opinions |
|  | in3 | Everyone in [activity1] made me feel wanted and accepted. |
|  | in4 | I felt like an important member of [activity1]. |
|  | in5 | I contributed to the choices made in [activity1]. |
|  | in6 | I felt safe in [activity1]. |
|  | in7 | I felt close to others in [activity1]. |
|  | in8 | I was satisfied with how much I got to help make decisions in [activity1]. |
|  | in9 | I felt a sense of shared purpose in [activity1]. |
|  | in10 | Being part of [activity1] became a big part of what kind of people we were. |
| ***Exclusion*** |  |  |
|  | ex1 | I felt isolated in [activity1]. |
|  | ex2 | I felt left out in [activity1]. |
| ***Positive interdependence*** |  |  |
|  | pi1 | My success in [activity1] depended on how well my whole group did. |
|  | pi2 | If important people saw the group doing well, they saw me doing well in [activity1]. |
|  | pi3 | Each person's task mostly depended on others in [activity1] doing their part. |
|  | pi4 | In the end, we were judged as a group/team instead of as individuals in [activity1]. |
|  | pi5 | Almost everything I did in [activity1] was to help my group reach its goal. |
|  | pi6 | It was important to me to respect decisions made by the group in [activity1]. |
|  | pi7 | I would give up what I want to help my group in [activity1]. |
| ***Negative interdependence*** |  |  |

|  |  |
| --- | --- |
| ni1 | Leaders in [activity1] encouraged us to compete against each other. |
| ni2 | People in the group/team competed against each other to see who was best in [activity1]. |
| ni3 | How often did everyone compete against each other in [activity1]? |
| ni4 | How often did you or your group compete against other groups in [activity1]? |
| ni5 | A goal of [activity1] was to win against other groups/teams of competitors. |
| ni6 | I felt like people were comparing me to others and judging me in [activity1]. |

**S5 Fig.**
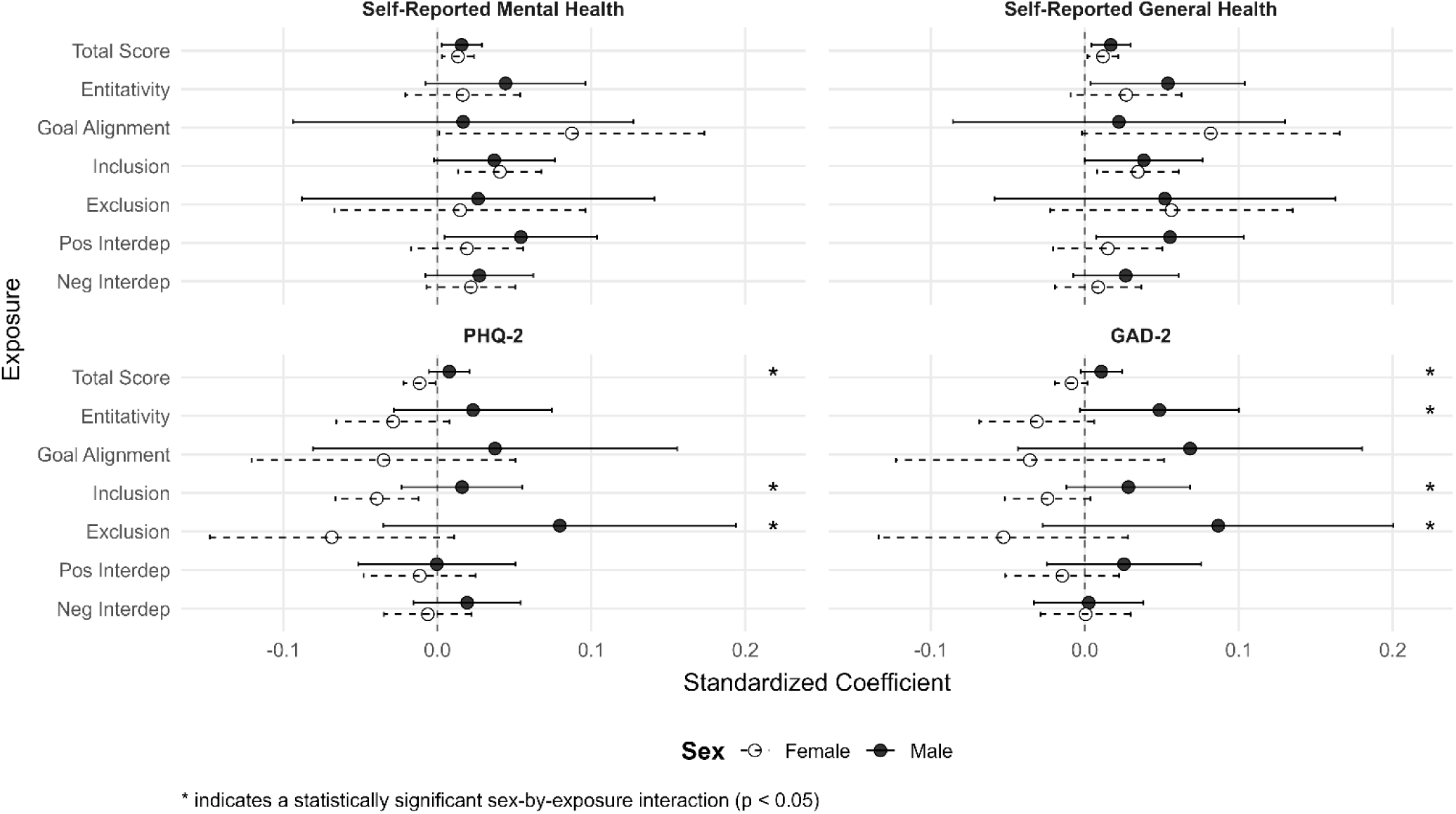
Sex-specific associations between cooperative experiences measure subscales and health indicators. Each panel represents a separately modeled health indicator (self-reported mental health, self-reported general health, PHQ-2, GAD-2). Cooperative experience measures (total; entitativity; goal alignment; inclusion and shared purpose; social exclusion; positive interdependence; negative interdependence subscales) are listed on the vertical axis. Circles represent standardized beta coefficients for each model (standard deviation change in the outcome per standard deviation change in the independent variable), and horizontal brackets represent 95% confidence intervals around the estimate. Hollow circles with dashed error bars correspond to female. Filled circles and solid error bars correspond to male. Asterisks indicate a statistically significant sex-by-exposure interaction (p < 0.05), denoting that slopes differ between sexes. All models were adjusted for age, Latino ethnicity, student status, and employment status.

